# Dengue seroprevalence study during COVID-19 pandemic in Bali

**DOI:** 10.1101/2022.07.12.22277538

**Authors:** Sri Masyeni, Rois Muqsith Fatawy, AAAL Paramasatiari, Ananda Maheraditya, Ratna Kartika Dewi, NW Winianti, Agus Santosa, Marta Setiabudy, Nyoman Trisna Sumadewi, Sianny Herawati

**Affiliations:** Department of Internal Medicine, Faculty of Medicine and Health Science, University of Warmadewa, Jl. Terompong 24, Denpasar, Bali, Indonesia; Infectious Disease and Immunology Research Center, Indonesia Medical Education and Research Institute, Faculty of Medicine, Universitas Indonesia, Jl. Salemba 6, Jakarta, Indonesia; Faculty of Medicine and Health Science, University of Warmadewa, Jl. Terompong 24, Denpasar, Bali, Indonesia; Faculty of Medicine, University of Udayana, Jl. Sudirman Denpasar, Bali, Indonesia

**Keywords:** DENV IgG antibodies, seroprevalence, infection

## Abstract

**Introduction:** Dengue infection poses significant public health problems in tropical and subtropical regions all over the world. The clinical manifestation of dengue varies from asymptomatic cases to severe dengue manifestation. The detection of clinical cases enables us to measure the incidence of dengue infection, whereas serological surveys give insights into the prevalence of infection. This study aimed to determine the dengue prevalence among healthy adult patients in Bali.

**Method:** Cross-sectional seroprevalence surveys were performed from July 2020 to June 2021 among healthy and adult patients in Denpasar Bali. Blood samples were collected from 539 randomly selected samples from urban sites in Denpasar. IgG antibodies against DENV were detected in serum using a commercial enzyme-linked immunosorbent assay (ELISA) kit.

**Results:** Overall, the positive dengue seroprevalence rate among 539 clinically healthy adult patients was high (85.5%). The subject’s median age was 34.1 (range between 18-86.1) years old. Most participants in the study were younger than 40 years old (61.2%). The gender is dominated by males (54.5%). The study found a significant association of dengue seropositivity among people age more than 40 years old with healthy status (*p*=0.005 and p<0.001, respectively). Another seroprevalence study reported a lower rate of dengue infection in children in Indonesia (69.4%). The difference may be associated with less probability of Aedes bites among the children. The study reflected the proportion of asymptomatic dengue that needs better assessment with a serological test.

**Conclusion:** The current study highlighted a high prevalence of dengue seropositive with a relatively dominant proportion of asymptomatic cases. The study guides the physicians o to beware of every dengue infection in tropical countries and prevents the spread of the disease.

## Introduction

Dengue has been considered the disease of the forthcoming, with alarming epidemiological patterns for both human health and the global economy[1]. The prevalence of dengue virus (DENV) infection has been predicted to cause 390 million new cases arising each year and approximately 96 million dengue hemorrhagic fever (DHF) cases leading to hospitalization each year[2]. Four serotypes of DENV (DENV-1, -2, -3, -4) have spread rapidly within countries and across regions, causing epidemics and severe dengue disease, hyperendemicity of multiple dengue virus serotypes in tropical countries, and autochthonous transmission in Europe and the USA[3,4]. However, dengue as well as other infections have been reported to have decreased during the pandemic[5,6].

The burden of dengue disease in many dengue-endemic countries including Indonesia remains poorly enumerated since the current passive surveillance systems capture only a trivial fraction of all dengue cases. Furthermore, it mostly depends on clinical diagnoses which exclude milder and atypical disease presentations [2,7]. Dengue transmission in Indonesia including Bali has become hyperendemic and a major public health problem. Dengue fever has spread throughout Indonesia since the first dengue case was reported in Surabaya in 1968 [8]. All four dengue serotypes have been reported from Indonesian regions such as Western Java, Surabaya, Bali, and Jambi. [3,9–13].

Dengue infection in Bali has become a recurring and significant public health issue. The study objective is to assess the prevalence of dengue infection detected by Indirect IgG ELISA.

## Material and methods

Cross-sectional seroprevalence surveys performed in July 2020 – June 2021 among healthy adults and non-dengue patients in Denpasar Bali. Blood samples collected from each of 539 randomly selected samples from urban sites in Denpasar. DENV IgG antibodies were checked in serum. A blood sample of 3 mL venous blood was taken into clot activator tubes from each participant. Serum was separated by centrifugation at 1300 ×g for 10 min, kept at +4 ?C, and transported to the laboratory of Universitas Warmadewa on the days for further processing.

DENV anti-IgG antibody detection in the collected sera was performed using a commercial enzyme-linked immunosorbent assay (ELISA) (catalog numbers EI 266a-9601-1 G, Panbio, Luebeck, Germany) following the manufacturer’s instructions. The kit’s specificity and sensitivity are 0.988 (95% CI: 0.979–0.993) and 0.892 (95% CI: 0.879–0.903), respectively. Briefly, serum samples diluted 1:101 were added to microplates and optical densities (OD) measured with a microplate reader. Antibody values ≥20 relative units/mL were considered seropositive.

### Ethical considerations

The study protocol was approved by the Institutional Review Board of the Faculty of Medicine, Udayana University, Bali, Indonesia (Approval No: 485/UN.14.2/KEP/2020). All study participants were enrolled after providing written informed consent.

### Statistical analysis

The descriptive analysis was used to describe the seroprevalence of dengue by Indirect IgG ELISA. The Chi-square test was used to analyze the association between the variables.

## Results

The study found a high prevalence of dengue infection among the participant. The baseline characteristics of the participants were listed in table 1. The results show the high prevalence of seropositive dengue among the participants can be seen in table 2.

**Table 1.** Baseline characteristics of the participants (N:539)

| Variables | Frequency | Percentage (%) |
| --- | --- | --- |
| **Age** |  |  |
| Median (IQR) | 34.1 (18-86.1) |  |
| < 40 y.o | 330 | 61.2 |
| ≥ 40 y.o | 209 | 38.8 |
| **Gender** |  |  |
| Male | 294 | 54.5 |
| Female | 245 | 45.5 |
| **Indirect IgG Dengue** |  |  |
| Negative | 72 | 13.4 |
| Equivocal | 6 | 1.1 |
| Positive | 461 | 85.5 |
| **Participants status** |  |  |
| Healthy | 373 | 69.2 |
| Any illness | 166 | 37.8 |
| **Diagnosis (N: 166)** |  |  |
| COVID-19 | 43 | 25.9 |
| Febrile illness | 33 | 19.9 |
| Non febrile illness | 90 | 54.2 |

**Table 2.** Association between the variables

| Variable |  | Indirect IgG ELISA |  |  |  | *p* |
| --- | --- | --- | --- | --- | --- | --- |
|  |  | Positif |  | Negatif |  |  |
|  |  | N | % | N | % |  |
| Age |  |  |  |  |  |  |
| < 40 y.o | 330 | 271 | 82.1 | 59 | 17.9 | 0.005 |
| ≥ 40 y.o | 209 | 190 | 90.9 | 19 | 9.1 |  |
| Gender |  |  |  |  |  |  |
| Male | 294 | 252 | 85.7 | 42 | 14.3 | 0.893 |
| Female | 245 | 209 | 85.3 | 36 | 14.7 |  |
| Participants status |  |  |  |  |  |  |
| Healthy | 373 | 336 | 90.1 | 37 | 9.9 | <0.001 |
| Any illness | 166 | 125 | 75.3 | 41 | 24.7 |  |

**Abbreviations:** IgG, Immunoglobulin G; DENV, Dengue Virus; ELISA, enzyme-linked immunosorbent assay; DHF, dengue hemorrhagic fever

## Discussion

The burden of dengue infection is not well documented in Indonesia, particularly among adult populations. The current dengue seroprevalence study conducted on adults in Bali reveals that people in tropical countries are at risk of dengue infection since the seroprevalence was found as high as 85.5%. Another seroprevalence study reports a lower rate of dengue infection in children in Indonesia (69.4%)[14]. The difference may associate with less probability of Aedes bite among the children than in the adult population. This study result is in line with another study that report the seroprevalence rate was high (91.5%) across all eight study sites. The reason is the study was conducted on adults and children in urban and rural populations in Thailand [15]. Study in Dhaka reports the seroprevalence as high as 80%[16]. Carabali et al. (2017) report the seroprevalence of dengue infection is 61% and only 3.3% of the seropositive subject has a past dengue history [17].

The current study also found that participants aged more than 40 years old had a significantly higher proportion of dengue seropositive than participants aged less than 40 (*p*=0.005). This finding may be related to the risk of Aedes bites being higher than younger age in their life. This is in line with another study report in Thailand, which found more than 98% of participants older than 25 years were found to be dengue seropositive[18].

Since the result history of suspected dengue infection was previously reported in only 60% of the participant, we predict that there was an asymptomatic dengue infection among the participant, compare with the results of the Indirect IgG Dengue ELISA result. The study results reflect the proportion of asymptomatic dengue that needs to assess with the serological study. Having this high seroprevalence (85.5%), the population have higher chance to be infected by dengue multiple times. Being exposed to this virus multiple times could increase the risk of severe disease [19,20]. An individual who is infected with a different dengue serotype for the second time may suffer antibody-dependent enhancement of infection, which means that antibodies from the previous infection serve to disseminate viral infection and increase viremia[19]. Therefore, the possibility of having high hospitalized patients of dengue infections should be of concern of the public health office in Bali.

The limitation of the study is the recall bias related to dengue infection history among the participants. Since we were only using Indirect IgG ELISA, we were unable to distinguish between primary and secondary infections. On the other hand, the IgG antibodies of dengue virus remain on the body for more than 5 years after the infection and the infection from years ago maybe detected on this study. Indeed, the cross-reactivity among the Flaviviruses infection needs to determine with the PRNT study that is not final yet in this study.

## Conclusion

The current study highlighted a high prevalence of dengue seropositive people with a relatively high proportion of asymptomatic dengue cases. The study results enable to guide the physician of the risk of more severe dengue in every dengue infection in tropical countries.

## Data Availability

All relevant data are within the manuscript and its Supporting Information files.

## Acknowledgments

We would like to thank all the participants, nurses, and the research team in the Faculty of Medicine and Health Sciences, Universitas Warmadewa for supporting this study.

## Authors contributions

Conceptualization: SM; data curation: AAALP, AM, RKD; formal analysis: NWW, AS, NTS; funding acquisition: SM; laboratory investigator: MS, SH; writing original draft: SM, RMF; writing-review & editing MS, RMF.

